# Challenges facing Canadian Long-Term Care Homes and Retirement Homes during the COVID-19 pandemic

**DOI:** 10.1101/2024.06.19.24308949

**Authors:** Christine Fahim, Ayaat T. Hassan, Keelia Quinn de Launay, Alyson Takaoka, Elikem Togo, Lisa Strifler, Vanessa Bach, Nimitha Paul, Ana Mrazovac, Jessica Firman, Vincenza Gruppuso, Jamie M. Boyd, Sharon Straus

## Abstract

COVID-19 presented a crisis for long-term care homes (LTCHs) and retirement homes (RHs). This study explored the pandemic-related challenges LTCHs and RHs faced and the strategies they used to mitigate them. Ninety-one key informant interviews were conducted with LTCH and RH leadership across 47 homes (33 LTCHs, 14 RHs) in Ontario, Canada from February 2021 to July 2022. Findings confirmed evidence for three main challenges. First, leaders were challenged to implement infection prevention and control protocols and measures. Second, they needed supports to facilitate COVID-19 vaccine access and to promote vaccine confidence. Third, LTCH/RH staff experienced significant well-being challenges in the face of COVID-19 pressures. Findings also reveal a plethora of strategies implemented by homes, with ranging reports of perceived success. Homes’ needs evolved rapidly as the COVID-19 pandemic progressed. The use of a co-creation, responsive and tailored approach to address evolving barriers and meaningfully support homes during emergencies is recommended.

**Key points:**

- COVID-19 challenges in homes persisted over one year into the pandemic
- We describe the IPAC, vaccine and wellness challenges faced by LTCH and RH
- We used these data to design a congregate care home support program to navigate COVID-19 challenges

## Introduction

The COVID-19 pandemic created a crisis in Canadian long-term care homes (LTCH) and retirement homes (RH). The first case of COVID-19 in Canada was confirmed on January 25, 2020; by March 2020 COVID-19 was declared a pandemic by the World Health Organization (Urrutia et al., 2021).

Canada has 2076 LTCHs, of which 627 (over 30%) are located in Ontario (Canadian Institute for Health Information [CIHI], 2021b). LTCHs offer a range of health and personal care services to individuals (primarily older adults) living in these assisted living dwellings, and these residents represent a population who are more susceptible to viral infection due to their congregate living arrangements and complex health care needs (Stall et al., 2020).

In the spring of 2020, >75% of COVID-related deaths in Canada were associated with LTCH and RHs (Public Health Ontario [PHO], 2020b). By July 7, 2020 there were more than 18,000 cases and 6851 deaths among residents of LTC and RH - the majority of cases and deaths being in LTC (Canadian Foundation for Healthcare Improvement [CFHI] & Canadian Patient Safety Institute [CPSI], 2020; Costa et al., 2021). One study found that Ontario residents living in LTC had a 13-fold increase in risk of COVID-19 related death compared to their community-living counterparts aged 69 or older (Fisman et al., 2020). During this period, LTCH staff infected with COVID-19 represented more than 10% of the country’s total cases (CIHI, 2020) and by December 2021, LTCH/RH residents accounted for 3% of Canada’s COVID-19 cases, but comprised 43% of Canada’s COVID-19 deaths (CFHI & CPSI, 2021; CIHI, 2021a).

Nearly one year into the pandemic, approximately 35% of Ontario’s LTCHs had experienced COVID-19 outbreaks (Stall et al., 2021; CIHI, 2021a).

The COVID-19 pandemic exposed a fragmented, under-resourced LTC system in Canada, with many long-standing failures that have been previously highlighted in more than 100 reports and public inquiries spanning back more than five decades coming to light (Estabrooks et al., 2023). Such failures include inadequate staffing and supports for staff, insufficient funding particularly in light of Canada’s aging population, a lack of standardized regulations that acknowledges the whole system, lack of data to inform quality improvement efforts, lack of involvement of staff, resident and family voices in the development of supports, and a need to address these gaps using a health equity lens in order not to exacerbate existing inequities (Estabrooks et al., 2020). Some, but not all of these issues apply to RHs as well, due to differences in regulation and oversight between the two settings (CFHI & CPSI, 2021). Efforts to rectify these systemic gaps in LTC persistent for over five decades remain limited, despite an abundance of high-quality evidence on effective solutions (Armstrong & Cohen, 2020; Estabrooks et al., 2023). Increasing LTCH resident complexity only further complicates this crisis in LTC – more and more residents entering LTCHs are functionally dependent with more complex health and social needs and advanced dementia (Estabrooks et al., 2023). With national failures evident and a lack of adequate support, education and renumeration provided for LTC staff even pre-pandemic, the LTC sector was simply unable to absorb the shockwaves of the COVID-19 pandemic and the sector’s lack of pandemic preparation plans (such as care plans for infected residents, access to personal protective equipment (PPE), lack of infection prevention and control (IPAC) training) exacerbated the crisis further (Estabrooks et al., 2023).

In the initial months of COVID-19, Canada experienced a far higher proportion of its COVID-19 deaths coming from LTCHs; these conditions were especially pronounced in the province of Ontario due to coordination gaps between pandemic preparedness, funding shortages, and insufficient staffing mix (Liu et al., 2020). Given the urgent need to support LTCH/RHs with outbreak prevention and management, our team members led a rapid review that informed the WHO guidance document on COVID-19 infection, prevention and control (IPAC) in LTCH (World Health Organization, 2021; Rios et al., 2020). Shortly after this, the Ontario Ministry of Health mandated that acute care hospitals should support LTCH to implement IPAC protocols and mitigate outbreaks in alignment with this evidence (PHO, 2020a; Ontario Ministry of Long-Term Care, 2022). While not included in the mandate, RH were also encouraged to adhere to these protocols. However, LTCH and RH leaders were challenged with implementing these IPAC protocols amidst a rapidly-evolving pandemic. It was in this context that we first aimed to design a responsive, co-created support program for Ontario LTCH and RH to help the staff in these settings navigate through their COVID-19 challenges.

To inform the components of our program to support LTCH and RH to navigate the COVID-19 pandemic, we conducted robust needs assessments with LTCH and RH in the province of Ontario - one of the Canadian provinces hardest-hit by COVID-19 (Government of Canada, 2023b). Needs assessments are useful tools for gathering information about a particular population, with the purpose being to bring about change beneficial to the health of said population (Stevens & Gillam, 1998). Our specific objectives were to: (1) explore the challenges faced by LTCH and RH leadership staff to navigate the pandemic safely and to identify resources that could assist them; and, (2) to determine what types of meaningful supports would strengthen their roles as leaders and their ability to guide their residents and staff through a pandemic. In this article, we present the findings of our needs assessments, which were conducted using key informant interviews with LTCH and RH leaders.

## Methods

We report our study findings using the Consolidated criteria for reporting qualitative research (COREQ). We conducted qualitative needs assessments using the Framework Method (Gale et al., 2013).

### Long-Term Care Homes and Retirement Homes Immunity Study

Given our extensive partnerships with LTCH and RH, we were approached by multidisciplinary colleagues to conduct immunity studies among LTCH and RH populations to assess SARS-CoV-2 seroprevalence and correlates of infection during the COVID-19 pandemic (Protocol: https://osf.io/wqrst). During enrollment, we quickly realized that while homes facing the COVID-19 crisis saw the benefit of the research, they had little bandwidth to contribute to it. In an effort to support these homes and facilitate this critical research, we proposed the development of a support program to be delivered alongside the immunity research (COVID-19 Immunity Task Force, 2021). We sought to use co-creation methods to identify and address key challenges facing LTCH and RH and use implementation science methods to iteratively adapt, implement and evaluate a support program that addressed these challenges. Thus, we approached homes with two yoked study requests. The first was to request enrollment in a COVID-19 immunity study that involved the collection of serosamples, dried blood spots, and wastewater data to track the prevalence, spread and correlates of infection and protection of SARS-CoV-2 among their staff, staff household members, LTCH/RH residents, and resident’s care partners/family members. The second was to request home leaders to participate in the design and implementation of a responsive support program that would provide tailored resources to navigate the pandemic.

### Study Design

We conducted semi-structured needs assessment interviews with LTCH and RH leaders to explore pandemic-related challenges, experiences implementing strategies to address these challenges, and needed supports. In keeping with other qualitative studies conducted during complex health emergencies, we used a rapid analysis approach to conduct and analyze our data in order to facilitate timely development of the support program (Johnson & Vindrola-Padros, 2017).

### Sampling and Setting

Our study was conducted in the province of Ontario (population 15.5 million), Canada. The Greater Toronto Area (GTA) is the most densely populated area in Ontario (population >6.2 million) (Statistics Canada, 2022) and was home to many “COVID hotspots” in the province (Jüni et al., 2021; Xia et al., 2022). Outbreaks in these hotspots were correlated with social determinants of health including income, level of education, higher proportions of visible minorities, housing density, and occupation (Xia et al., 2022; Nadesan, 2022). We primarily sampled homes in the GTA and its surrounding regions; a minority of homes were in the Ottawa and Champlain regions. Homes were eligible to participate in our study if leaders (1) provided consent to participating in both the immunity and implementation research, (2) were located in Ontario, and (3) provided a designated communication ‘point person’ within the home. Homes unable to meet these criteria were excluded from the study, as were Indigenous LTCH and RH given that our study team did not have the appropriate expertise and resources to meaningfully support Indigenous partnerships.

### Participant Recruitment

We used a variety of passive and active recruitment strategies to enroll homes in our study; study advertisements were shared using study websites, our project partners (including a number of LTCH, RH, patient advocacy, government and research organizations), social media. Project partners and participants were also encouraged to share the advertisements with their networks. Within each home, we recruited LTCH and RH leadership staff (defined as decision-makers responsible for day-to-day operations in an individual LTCH or RH) to participate in key informant interviews. Leaders were eligible to participate in an interview if they were at least 18 years of age, were comfortable speaking English or French, and agreed to provide individual consent for participation. Only one leader per home was required for these interviews, however, participants were given the option of participating in group interviews.

### Participant Characteristics

Table 1**: Demographic Characteristics of Homes.** We conducted 91 key informant interviews with LTCH or RH leadership staff across 47 homes (33 LTCH and 14 RH) between February 2021 and July 2022. Our sample included privately owned for-profit homes (63.8%), as well as privately owned non-profit (17.0%), and publicly owned, non-profit homes (19.2%) (Table 1).

**Table 1:**
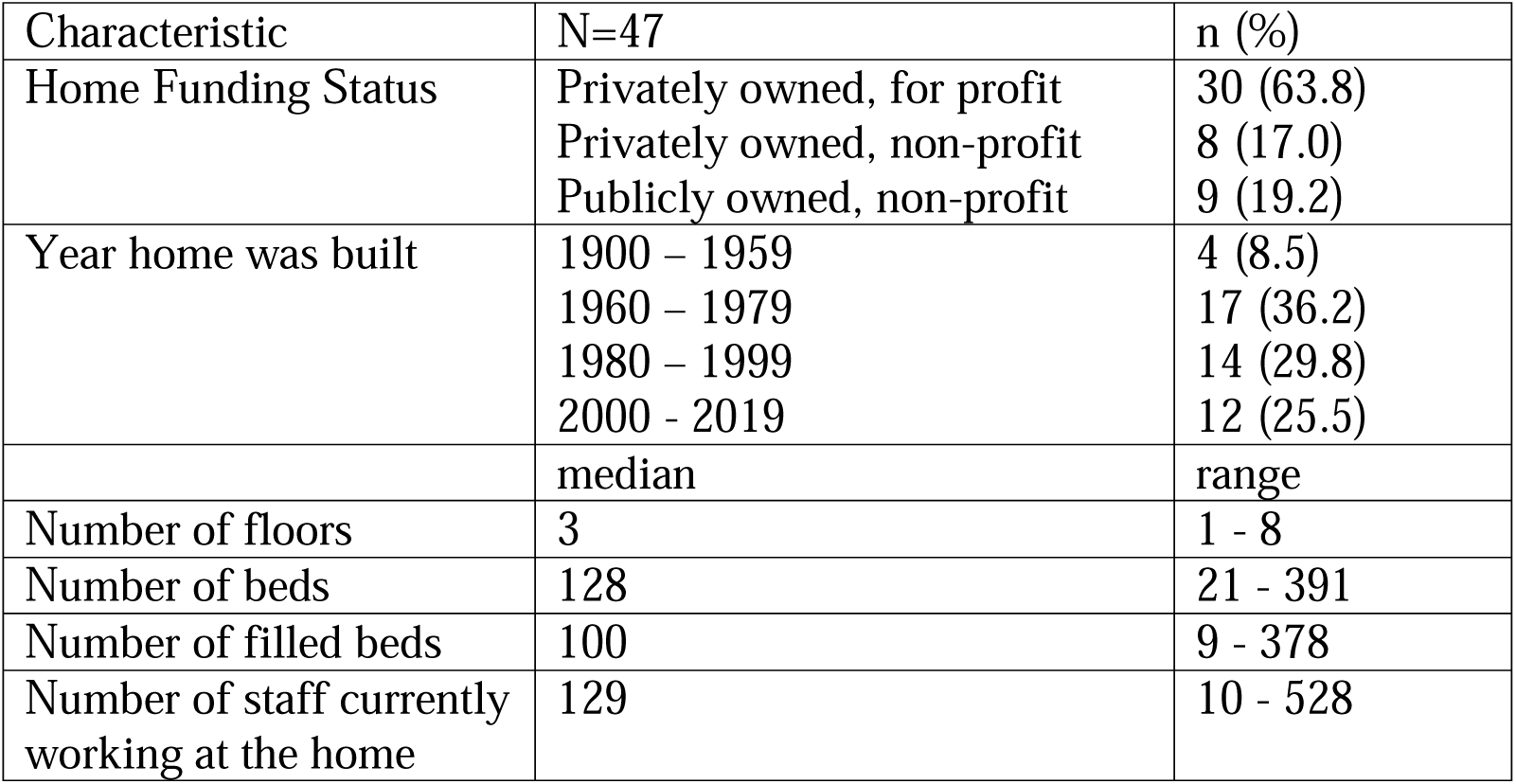
Demographic Characteristics of Homes.

| Characteristic | N=47 | n (%) |
| --- | --- | --- |
| Home Funding Status | Privately owned, for profit<br>Privately owned, non-profit<br>Publicly owned, non-profit | 30 (63.8)<br>8 (17.0)<br>9 (19.2) |
| Year home was built | 1900 – 1959<br>1960 – 1979<br>1980 – 1999<br>2000 - 2019 | 4 (8.5)<br>17 (36.2)<br>14 (29.8)<br>12 (25.5) |
|  | median | range |
| Number of floors | 3 | 1 - 8 |
| Number of beds | 128 | 21 - 391 |
| Number of filled beds | 100 | 9 - 378 |
| Number of staff currently working at the home | 129 | 10 - 528 |

The median number of floors in each home was three (range 1-8) with 128 beds (range 21-391) overall, and 129 staff per home (range 10-528). The majority of the homes were located in the Greater Toronto and Hamilton areas (n=35).

### Data Collection

In keeping with an integrated KT approach, we co-designed an interview guide with LTCH/RH stakeholders (Canadian Institutes of Health Research, 2015). While the initial focus of the guide was to investigate IPAC implementation challenges, stakeholders highlighted two other key areas of focus. First, stakeholders described the need to probe about challenges related to COVID-19 vaccines and second, stakeholders described the physical, emotional and mental toll the COVID-19 pandemic had on LTCH/RH staff. In response, we created an interview guide that included probes to investigate IPAC, Vaccine and Wellness challenges. To ensure we were not leading participants during the interviews, we also invited participants to generally describe their experiences during the COVID-19 pandemic, to list any challenges they experienced challenges with, and to describe strategies they used or needed to address these challenges.

Data were collected between February 2021 and July 2022. Participants took part in a 20- to 25-minute semi-structured interview (Appendix B) via phone or Zoom (https://zoom.us/) conducted by an interviewer (AT, KQD, AH, JF, MSc or MPH education) and note-taker (AM, VB, OS, BSc or BPH education). Verbal consent and interviews were recorded. Notes were taken as close to verbatim as possible, in keeping with rapid analysis methodology (Nevedal et al., 2021). After the data collection session, the interviewer and note-taker reviewed the notes for accuracy; the note-takers listened to the recordings to supplement any missing details in the notes (Nevedal et al., 2021). All participants were de-identified and assigned a unique study ID. Participants were not compensated.

### Characteristics of Interviewers

Interviewers were Research Coordinators from the Knowledge Translation Program at St. Michael’s Hospital. They were all women and had experience in qualitative research, implementation, or community-based participatory approach research. The research staff were not known to the study participants. Our research team recognizes that positionality is intersecting and fluid; our team shares some commonalities (e.g., gender), while other characteristics and viewpoints differ. With respect to this project, our team included individuals who work with older adults as clinicians, researchers, caregivers, and those with experiences of loved ones in long-term and retirement care. Our approach stemmed from a co-creation and integrated knowledge translation lens (Jull et al., 2017) that could allow space for those working in LTCH and RH to identify their needs, which would later inform the development of relevant, tailored interventions.

### Data Analysis

Using a rapid analysis approach (Nevedal et al., 2021), data were double-coded by research staff and trainees experienced in implementation science methodology (KQD, LS, ET, AH, JF). Five transcripts were double-coded using open coding to generate a codebook. This codebook was used to categorize the challenges homes faced and strategies implemented to address these challenges. Experienced research staff double coded 15 interviews; discrepancies were resolved until there was 100% agreement. The remaining interviews were single-coded.

Data were then themed by two researchers (AH, MSc; LS, PhD) with input from a scientist (CF, PhD).

### Funding and Ethics

This study was funded by the COVID-19 Immunity Task Force and the John and Myrna Daniels Charitable Foundation via the University of Toronto’s Aging and Place Institute. This study received ethics approval from the Toronto Academic Health Science Network (REB 20- 347).

## Results

We categorized the identified challenges within the overarching themes identified by our LTCH/RH partners. Specifically, three themes were identified by our partners: implementation of IPAC protocols and measures; facilitation of access and uptake of SARS-CoV-2 vaccines amidst vaccine mandates, and well-being challenges, particularly among LTCH/RH staff. In

Tables 1-3, we report the challenges related to these three categories and describe the strategies used by home leadership to address them.

### Infection Prevention and Control

Table 2**: Infection Prevention and Control Challenges and Strategies Implemented.** We identified ten challenges related to IPAC implementation during the COVID-19 pandemic (Table 1). These included: insufficient resources and time to deliver IPAC education and preparation (e.g., mask fitting), lack of consistent IPAC implementation by staff and residents with limited capacity to follow protocols (e.g., proper masking), difficulties keeping up with rapidly evolving COVID-19 protocols and mandates, resource (including personal protective equipment [PPE] and COVID-19 rapid test) shortages, impact of the physical home structure on IPAC implementation (e.g., lack of physical space to cohort and distance staff and residents), family pushback on IPAC protocols, staff PPE fatigue, and fears of returning to normal and loosening IPAC restrictions. To address these challenges, homes commonly leveraged external supports from hospitals and public health units to receive updates on COVID-19 mandates, protocols and their implementation, in addition to physical (e.g., equipment), financial, and human resources. Homes found it useful to have a dedicated IPAC champion to provide advice, guidance and staff support. Some homes implemented multi-pronged strategies (e.g., huddles, use of champions, handouts, training) to facilitate IPAC uptake; others also implemented routine audits. Other levers to IPAC implementation included having leaders who were committed to transparent and open communication, leaders with previous experience managing health emergencies, and homes with physical space conducive to IPAC cohorting and isolation.

**Table 2:**
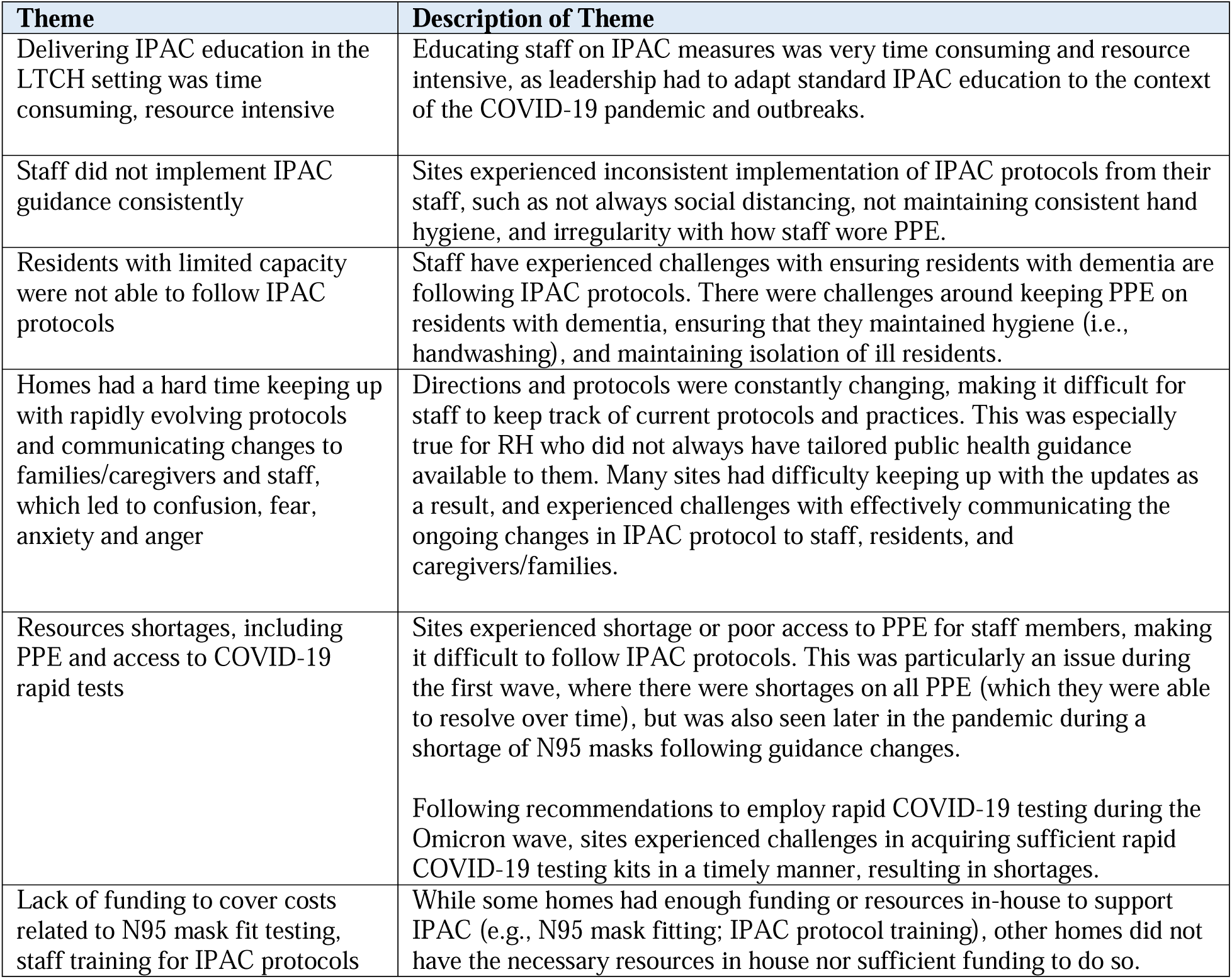
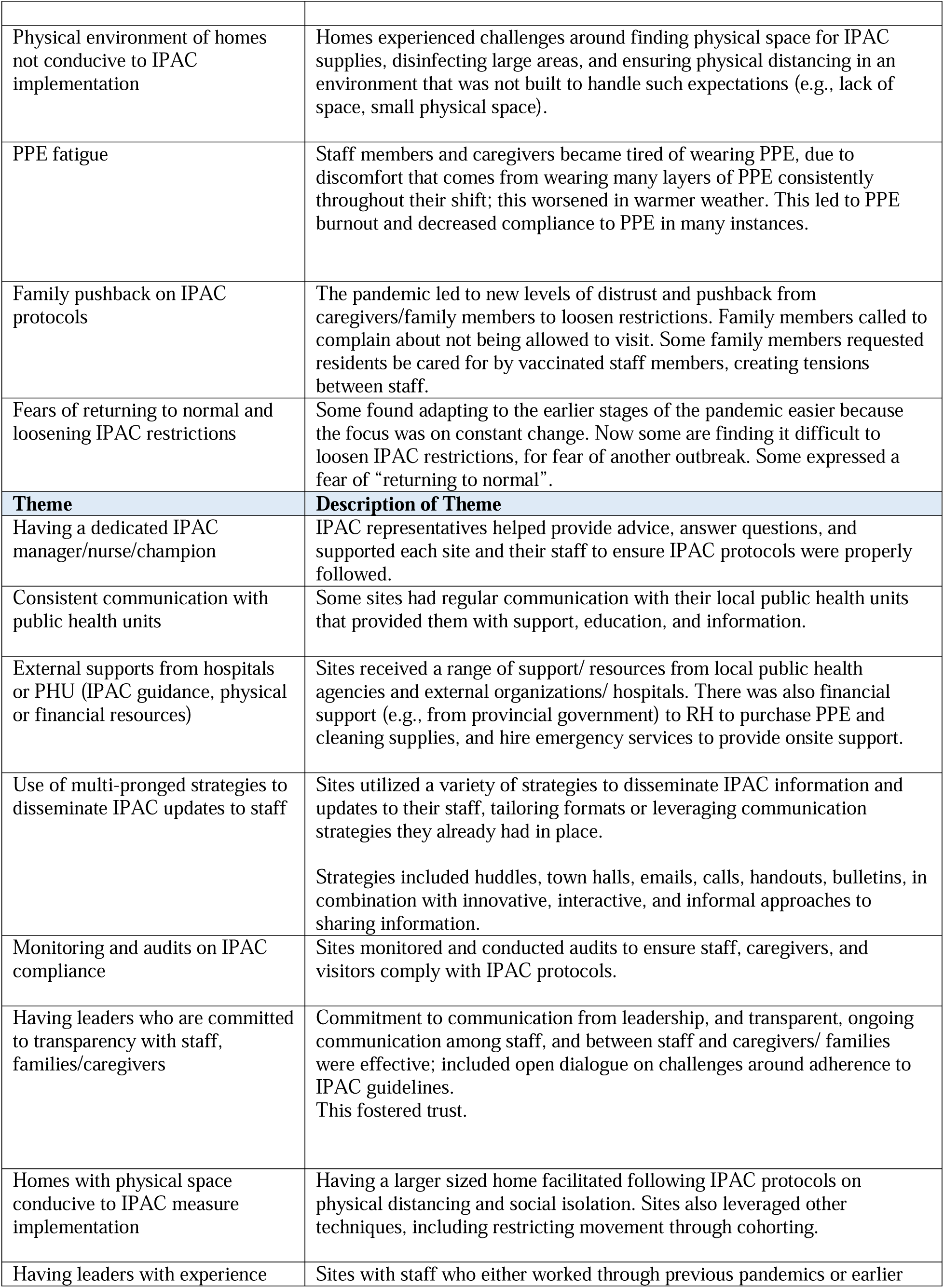
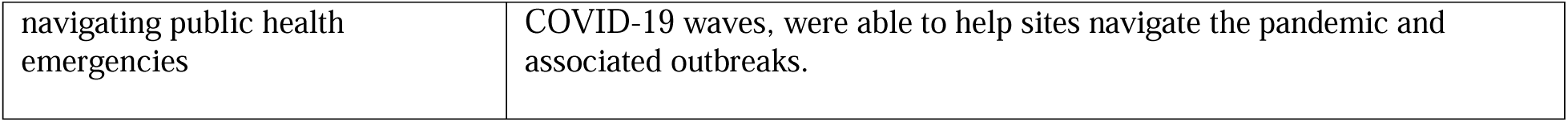
IPAC Challenges and Strategies Implemented.

### COVID-19 Vaccines

Table 3**: COVID-19 Vaccine Challenges and Strategies Implemented.** Notably, participants reported that the majority of LTCH and RH staff were supportive of COVID-19 vaccines. However, we identified six challenges to COVID-19 vaccine uptake in LTCH and RH (Table 2). Barriers at the individual staff level included mistrust around vaccine safety, beliefs that COVID-19 boosters would not improve health outcomes, and beliefs that vaccine mandates were an infringement on labour laws and personal liberties. Some staff did not feel comfortable working with residents or colleagues who were unvaccinated, which led to workplace conflict and tension. Furthermore, family members concerned about vaccine safety did not provide consent for LTCH residents to receive the vaccines.

**Table 3:**
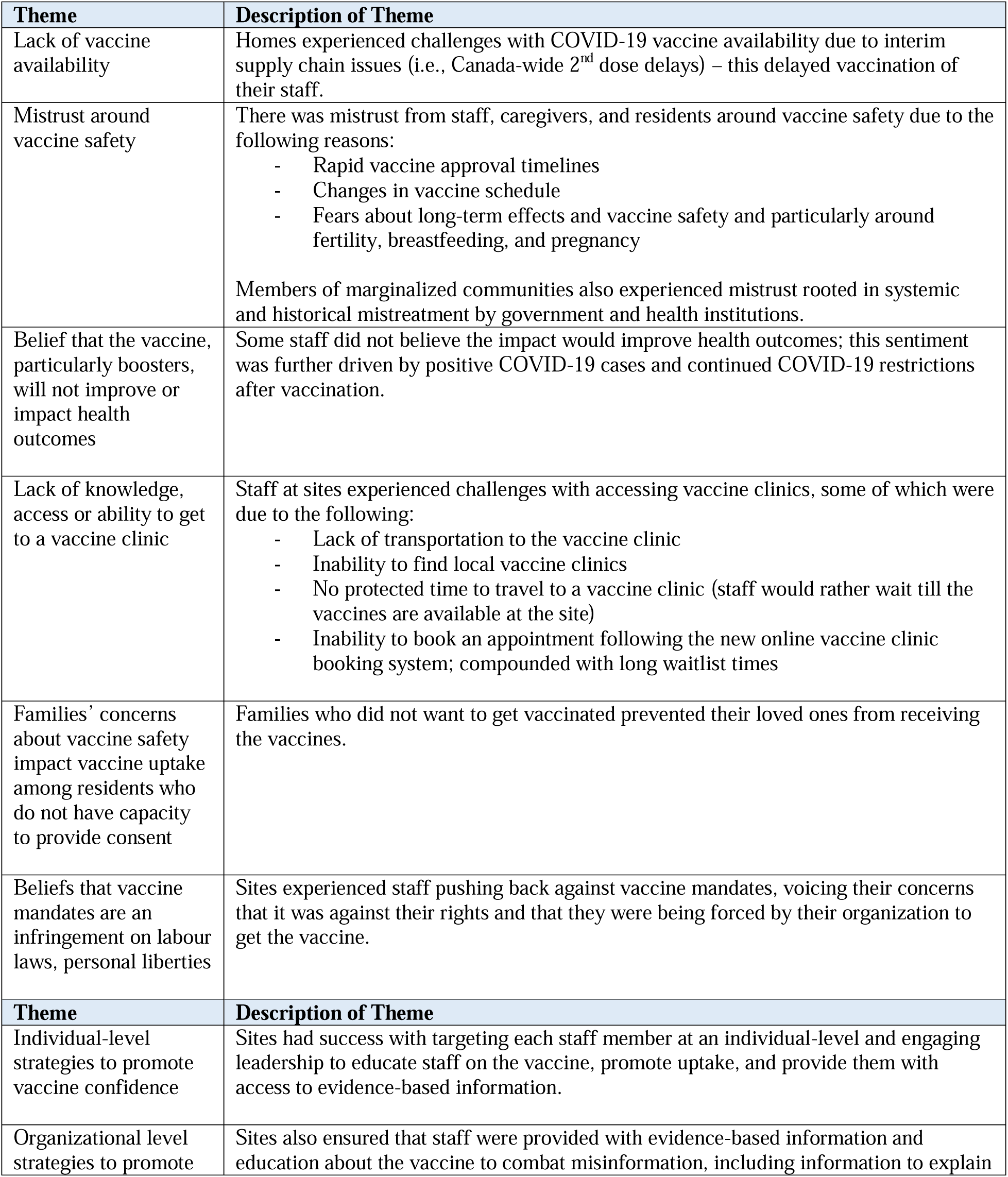
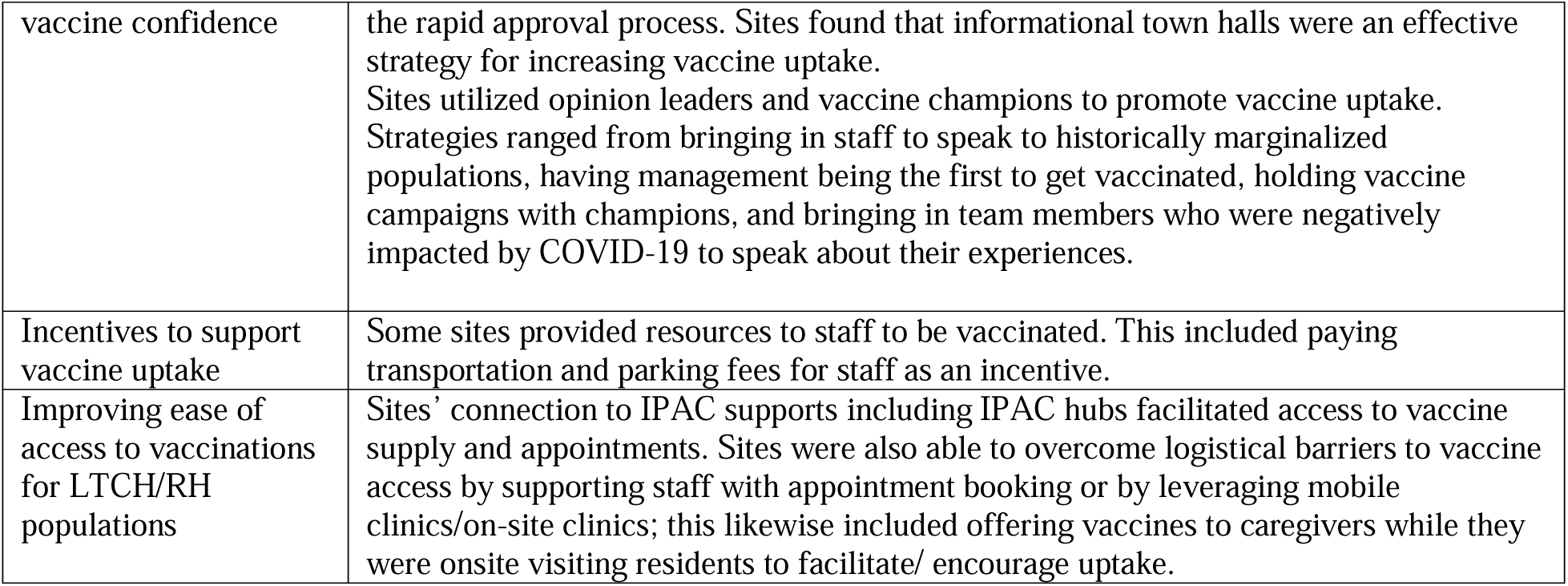
COVID-19 Vaccine Challenges.

Also reported were logistical barriers, including a lack of knowledge or ability to access a vaccine clinic, challenges using the online booking systems, and lack of vaccine availability due to Canada-wide supply chain issues. In particular, some RH leaders reported that their homes, unlike LTCH, were not prioritized to receive the COVID-19 vaccines and were challenged to advocate and secure doses for their staff members. Government-wide mandatory vaccination policies were perceived both as a barrier to uptake (some viewed it as an infringement on personal liberties) and a facilitator to uptake (led the majority of LTCH/RH residents and staff to receive the first two doses), though uptake of subsequent COVID-19 boosters remained a persistent challenge. Doubts about COVID-19 booster necessity was driven by beliefs that people had COVID-19 antibodies from their initial doses or from natural illness, and because COVID-19 case numbers were increasing despite high vaccination rates.

Enablers to vaccine uptake were also identified. Some staff believed that vaccine uptake (particularly the initial two doses) would facilitate a return to normalcy, for instance, by ending lockdowns and allowing staff to return to work in more than one home. Others became discouraged when public health mandates remained unchanged (e.g., lockdowns) and COVID-19 cases continued, following uptake of COVID-19 vaccines.

In other homes, outbreaks drove staff, who were initially hesitant, to receive the vaccines. Participants reported the use of multi-pronged strategies to address vaccine misinformation and concerns, including 1-to-1 conversations with staff, engaging leadership to provide education about the vaccines, distribution of educational materials to address key concerns, and holding town halls with respected experts. Use of opinion leaders and respected vaccine champions were also found to be effective strategies to combat vaccine hesitancy, particularly when champions included members of historically marginalized populations and reflected the demographics of the LTCH/RH staff population. LTCH/RH policies such as provision of financial resources (e.g., transportation, parking coverage) and providing incentives to vaccination were perceived as effective strategies. Finally, homes used connections to IPAC supports, including IPAC hubs that included regional hospitals, to facilitate access to vaccine supply and appointments, particularly at the start of the vaccine rollout. Some sites overcame logistical barriers to vaccine access by supporting online appointment booking or hosting mobile or on-site vaccine clinics. Some homes also offered boosters to caregivers while they were visiting residents.

### Staff Well-Being

Table 4**: Well-being Challenges and Strategies Implemented.** LTCH/RH leaders reported that frontline staff such as PSWs and nurses experienced a variety of challenges to well-being during the pandemic and described significant experiences of burnout, low morale, and mental health challenges. These were driven by working in a high-stress, high-risk environment coupled with challenges outside of the workplace, such as fear of transmitting COVID-19 to their families, colleagues and residents, childcare challenges, fears about vaccine safety, and general fears about the novel virus and its impacts (including health and economic impacts). Provincial policies aiming to curb COVID-19 spread prevented staff from working in more than one home, which led to staff shortages and increased burden on existing staff. Policies on social distancing prevented the implementation of social and activities programming for residents and increased resident isolation, which led to decreased social interaction and subsequent declines in residents’ physical and mental health. Participants reported that they and their staff felt helpless to address these concerns and were unable to support their residents’ needs.

**Table 4:**
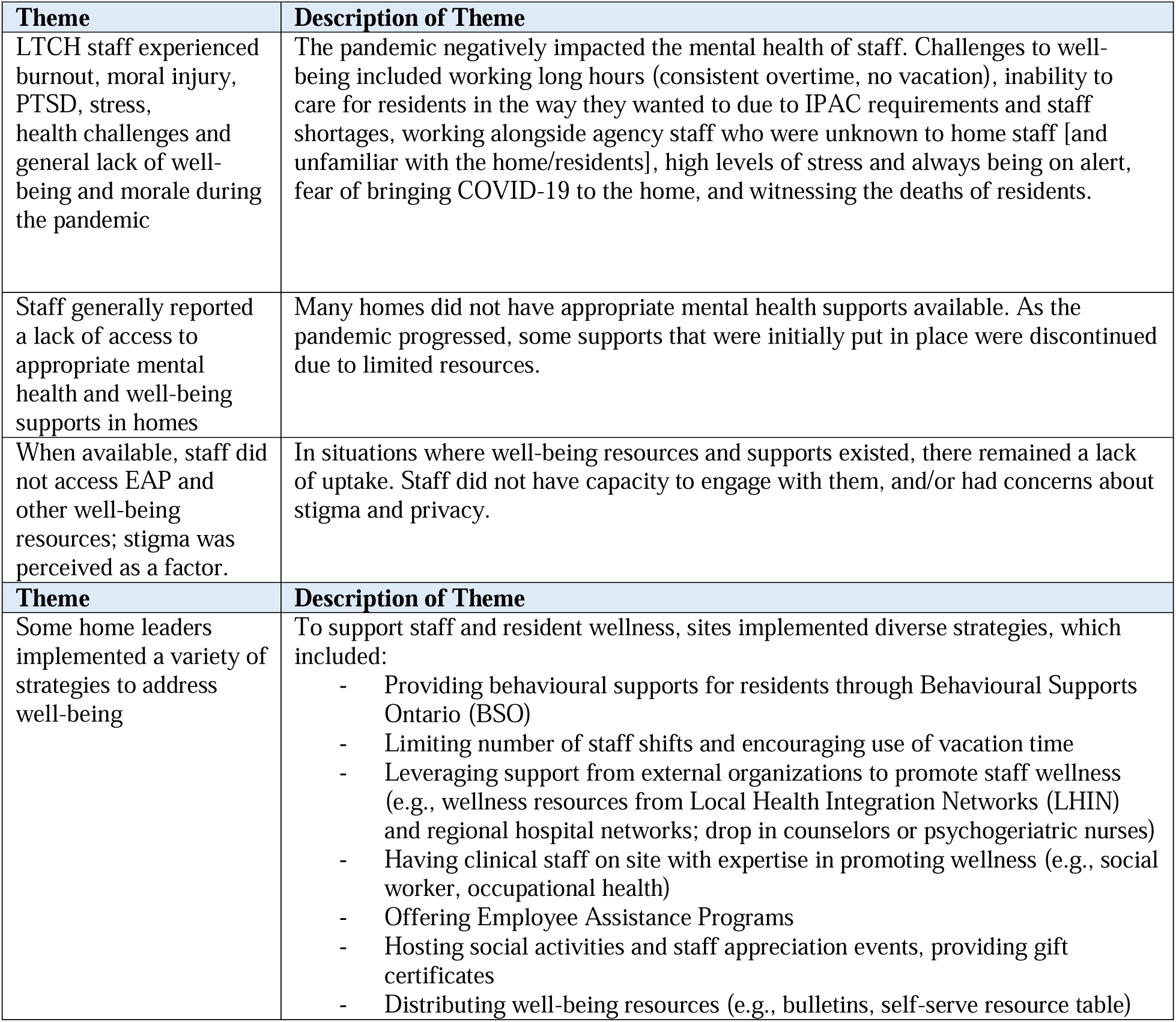
Well-being challenges and implemented strategies.

Staff also became sick with COVID-19, saw colleagues and family members become sick with COVID-19 and witnessed the death of residents. As the pandemic continued, staff were expected to maintain high rates of compliance with IPAC protocols, including wearing masks while much of the province had lifted masking requirements. This led some staff to experience IPAC fatigue, which was compounded by emotional and physical burnout. Some staff refused to work with residents who had COVID-19 and others left their positions or chose to retire early due to burnout and feelings of ‘moral injury’ (i.e., feeling unable to care for residents in an optimal way while feeling unsupported by leadership). In turn, these challenges increased staffing shortages and pressures on remaining staff. For instance, role shifting was prevalent with many frontline staff (and sometimes managers) taking on responsibilities outside of their traditional tasks (e.g., personal support workers implemented IPAC protocols, nurses became IPAC practitioners, and managers supported resident care) while many staff worked longer hours or double shifts to meet home and resident needs. When homes used agencies to address staff shortages, regular staff members worked alongside people they did not know or trust, which sometimes led to increased stress, workplace conflict, reduced staff morale and team cohesion.

LTCH/RH leadership also faced a number of unique challenges. In addition to trying to maintain home functioning and staff morale, leaders were required to participate in daily or weekly calls with external organizations such as public health units or hospital IPAC hubs. System inefficiencies also challenged home leaders; for instance, some participants reported being forced to individually source PPE amidst the country-wide shortages. One participant reported having to source equipment from three medical providers, as the government had not yet developed its centralized system. Leaders also struggled to stay current with continually changing COVID-19 mandates, policies and guidance, and found it challenging to address questions about the nature of the virus or, as the pandemic evolved, COVID-19 vaccines. Participants also reported high levels of distrust directed toward them from by staff and by residents’ family members who were frustrated by IPAC protocols and lockdowns which limited or eliminated visits to loved ones.

Leaders reported feeling insufficiently equipped to provide resources to staff to address these challenges. At the height of the pandemic, they prioritized implementation of IPAC protocols and resident care, which left little capacity to develop and implement wellness programs to support staff. Some did report the implementation of well-being activities in the workplace, however, they were also reports of discontinuation of these activities due to a lack of capacity or funding to sustain them. Other participants implemented and encouraged mental health supports such as employee assistance programs, but noted little uptake among staff due to accessibility challenges and stigma associated with seeking mental health supports.

Despite these challenges, leaders committed to staff well-being constantly found opportunities to implement multi-pronged and tailored strategies (see Table 3). Some leaders used a co-creation approach to develop and implement these strategies, such as the formation of a wellness committee or providing opportunities for leadership to listen to staff concerns.

## Discussion

The impact of COVID-19 on long-term care and retirement homes was devastating. LTCH/RH staff were at increased risk of becoming sick with COVID-19, not only through workplace exposures, but because they often resided in neighborhoods with lower household incomes, higher rates of ‘essential/frontline’ workers, and higher rates of COVID-19 (Ma et al., 2022). Nearly one year into the pandemic, we conducted 91 key informant interviews with LTCH and RH leadership in 47 homes to assess their experiences navigating the pandemic and to define the challenges faced by LTCH and RH.

Participants identified three overarching challenges associated with IPAC implementation, COVID-19 vaccine uptake, and overall staff well-being. In the early days of the pandemic, concerns were related primarily to a shortage of PPE resources (e.g., masks, face shields) and IPAC protocol implementation issues. Many of these challenges have been well-reported and highlighted as significant concerns for congregate homes for decades (Estabrooks et al., 2020; Estabrooks et al., 2023; Rochon et al., 2022). These challenges include physical infrastructure not conducive to cohorting, distancing or isolation, and limited resources to support the implementation of IPAC protocols (e.g., IPAC specialists) (Estabrooks et al., 2020; Estabrooks et al., 2023). Other contextual challenges include supply chain shortages (Healthcare Excellence Canada, 2020; Houston et al., 2023), which limited access not only to PPE, but also to COVID-19 PCR testing (when rapid antigen tests had not yet become widely available) (Sinha et al., 2021).

As PPE supplies became more available and IPAC protocols became routinized with the support of LTCH/RH leadership, hospital and public health units, other challenges emerged.

When COVID-19 vaccines entered our health system in December 2020, home leaders advocated for early access for their residents and staff. While this prioritization was later granted to LTCH, this was not the case for RH, creating equity imbalances (Sinha et al., 2021; Mishra et al., 2023; Estabrooks et al., 2023; Clark et al., 2023). Further, home leaders contended with staff and family fears about vaccine safety and efficacy; these concerns further escalated with the Ontario government’s introduction of mandatory vaccination policies in the spring of 2021 (Ontario Newsroom, 2021).

Frequent changes to IPAC and vaccination policies posed further challenges. In Canada, these included conflicting information about the safety of mixing vaccines, the safety of the AstraZeneca vaccine, vaccine schedules, and the speed at which vaccines were approved (AlShurman et al., 2023). The absence of clear rationales and mixed messaging created confusion and eroded trust among staff and families, fostering fear, anger, and resistance to policies like vaccine mandates, family visitation, and masking (Rochon et al., 2022). Multiple studies have demonstrated the link between inconsistent, unclear vaccination policies and a rise of vaccine hesitancy (Vernon-Wilson et al., 2023). These policies reinforced doubts about the vaccines and fueled mistrust of health organizations, especially among historically marginalized communities (Fahim et al., 2023b; Fahim et al., 2023a; Theivendrampillai et al., 2023). These sentiments extended to LTCH/RH and other healthcare staff, with high levels of vaccine hesitancy reported as the vaccines were first rolled out (Murmann et al., 2023). This context added significant strain on home leaders, who did not feel well equipped or supported to navigate access, uptake, and morale challenges (Vernon-Wilson et al., 2023; AlShurman et al., 2023; Lowe et al., 2022).

Challenges to staff well-being were consistently reported in our interviews. Frontline staff increasingly faced stress, resulting in PTSD from watching residents and colleagues become ill and die from COVID-19 while at the same time experienced burnout and other negative effects of short-staffing (Fisher et al., 2021). They were confronted by feelings of guilt, at times leading to moral injury (Reynolds et al., 2022) when they watched residents deteriorate from a lack of socialization or activity programming and visitations due to IPAC policies, as they lacked the power or authority to address this (Healthcare Excellence Canada, 2020). Studies have described LTCH/RH workers as the “forgotten front line” (Fisher et al, 2021). While other healthcare workers were being hailed as heroes, LTCH/RH staff, particularly staff in unregulated roles, such as Personal Support Workers, remained underpaid, underappreciated and under-supported (Mishra et al., 2020; Sultana & Ravanera, 2020; Sathiyamoorthy, 2020 ; Hapsari et al., 2022). These workers, who provide 90% of direct resident care, often work as part-time or casual employees in multiple homes. Mandates such as the ‘one-home policy’ requiring staff to only work in one home, coupled with province-wide shutdowns, directly impacted staff incomes and amplified needs for basic services such as childcare support and transportation (Reopening Ontario Act, 2020). Unsurprisingly, such challenges directly impacted well-being and mental health and compounded the issues staff faced at work. Racialized staff (estimated at >40% of the Ontario personal support worker workforce) also experienced intersecting challenges related to racism and workplace violence (Ontario Ministry of Long-Term Care, 2020; Sethi, 2020; Ejaz et al., 2011). In our study, we identified a few homes with robust employee support programs or supports; those that did have services available reported low uptake by staff due to lack of capacity to engage, lack of awareness or concerns about stigma.

Our participants described a plethora of strategies that were used to mitigate these challenges. With respect to IPAC and vaccines, homes found it helpful to appoint dedicated champions or facilitators to support compliance and uptake. Many homes reported developing educational resources and conducting audits to measure IPAC and vaccine compliance. Others praised their public health units who provided a range of supports to homes, from educational materials to equipment (e.g., PPE and cleaning supplies) to financial supports to homes. Leaders also described the benefits of the hub-and-spoke care delivery model (Government of Canada, 2023c), which was implemented to allow hospitals to ‘partner’ with LTCH and provide supports; notably, these integrations were largely absent pre-pandemic (Estabrooks et al., 2020; Estabrooks et al., 2023). Having effective leadership that was empathetic, collaborative, and engaged was consistently cited as a facilitator to combatting challenges related to IPAC, vaccines and well-being. In our study, participants reported implementing various strategies to maintain staff well-being and morale during the pandemic, including providing access to behavioral supports for residents, encouraging vacation time, offering employee assistance programs, and hosting wellness days and activities. Still, these were often deemed insufficient in the absence of government-wide policies such as paid sick leave for COVID-19 testing, vaccinations (Ontario Ministry of Labour, Immigration, Training, and Skills Development, 2023), or illness and financial benefits (Government of Canada, 2023a).

The COVID-19 pandemic put a spotlight on longstanding inequities and systems gaps for the LTCH and RH sectors and their workers in Canada. These factors had a direct impact on homes’ abilities to respond to novel coronavirus, resulting in a tragic crisis. Ontario’s Long-Term Care COVID-19 Commision published its final report in April 2021, noting that, “many of the challenges that had festered in the long-term care sector for decades- chronic underfunding, severe staffing shortages, outdated infrastructure and poor oversight – contributed to deadly consequences for Ontario’s most vulnerable citizens during the pandemic.” (Marrocco et al., 2021). Our findings are consistent with the well-documented challenges in LTCH and RH and other research conducted at this time (Stuart, 2022; Baumann et al., 2022; Ayalon et al., 2020; Syed & Ahmad, 2021; Hung et al., 2022; McGilton et al., 2021). However, our data also show that the systemic challenges reported by other researchers earlier in the pandemic continued to persist well through 2021 and 2022 despite massive media and government attention and efforts to address these challenges (Wong et al., 2021; Ayalon et al., 2020; Estabrooks et al., 2020). As described in *Restoring Trust* policy briefing, “COVID-19 [was] a shock wave that cracked wide all the fractures in our nursing home system” (Estabrooks et al., 2020, p. 8). Despite these systemic challenges, a number of factors were identified as facilitators to a strong response to COVID-19. In 2022, Baumann and colleagues found homes that were able to mobilize early had strong leaders that quickly implemented the required training and restructuring to implement Ministry of Health and Long-Term Care directives, and were directly involved in the response (i.e., on the ground with frontline staff, rather than in an office or working from home). Effective managers were those who provided clear, consistent and continuous communication to staff while minimizing over-communication to reduce cognitive overload. Systematic review data further identified home leadership quality and feeling supported as direct correlates to LTCH staff’s burnout and quality of life indicators (Costello et al., 2019).

Our study was one of few in Ontario that was conducted with the intent of designing immediate and sustained strategies to holistically support LTCH and RH leaders to navigate the pandemic and post-pandemic period (Zelmer et al., 2023; Glowinski et al., 2022). We leveraged these findings to design the Wellness Hub program, which we describe in detail elsewhere (https://wellness-hub.ca/). Briefly, Wellness Hub delivered educational resources and tailored strategies to support IPAC, vaccine and well-being concerns in LTCH and RH. Unlike other interventions with a singular focus (Glowinski et al., 2022), our program leveraged province-wide LTCH supports (e.g., Healthcare Excellence Canada’s LTC+ program) and aimed to deliver a comprehensive program with integrated supports for IPAC, vaccine confidence and staff well-being in order to streamline implementation and reduce burden (Zelmer et al., 2023). By iteratively collecting needs assessment data while designing our support program, we were able to be nimble, adaptive and responsive to the rapidly-changing COVID-19 context.

## Limitations

Our study has limitations. It was limited to homes in Ontario, Canada, and mostly included homes located in the Greater Toronto Area. We interviewed LTCH and RH leadership to quickly facilitate a needs assessment to inform the development of a subsequent support program; however, the perspectives of LTCH and RH residents, caregivers, and staff are important voices missing from our assessment. Finally, we conducted these interviews over a 17- month period between February 2021 and July 2022. Thus, reported challenges facing homes in the early waves of the pandemic (March 2020 – February 2021) may have been subject to a recall bias, though we anticipate the impact of this bias is limited, given consistency of our findings with other research reported during this time. Our findings demonstrate credibility, confirmability and we have extensively reported our methodology in order to enhance dependability. Our study setting was limited to LTCH and RH in the province of Ontario, mainly in the Greater Toronto Region. The transferability of our findings may be limited compared to other settings, including rural Ontario and other regions in Canada, although based on other national reports, we anticipate these challenges may have been similar, though perhaps experienced to different degrees, across homes in Canada (Stenfors et al., 2020).

## Conclusion

LTCH and RH experienced significant challenges during the pandemic related to IPAC implementation, uptake of COVID-19 vaccines and staff well-being and mental health. We recommend that evidence-based strategies should be identified and implemented using a co-created, tailored approach in order to mitigate these barriers and leverage strengths and facilitators. We used the findings of this study to develop a program to support long-term care and retirement homes to navigate the COVID-19 pandemic.

## Data Availability

All data produced in the present study are available upon reasonable request to the authors

## Acknowledgements

We would like to thank Chelsea Gao, Oswa Shafei, and Jane Dim for their contributions to data collection.

