## Supplemental Table 2 for "Challenges facing Canadian Long-Term Care Homes and Retirement Homes during the COVID-19 pandemic"

| **IPAC Challenges Experienced by LTCH/RH During the COVID-19 Pandemic** | | |
| --- | --- | --- |
| **Theme** | **Description of Theme** | **Quote** |
| Delivering IPAC education in the LTCH setting was time consuming, resource intensive | Educating staff on IPAC measures was very time consuming and resource intensive, as leadership had to adapt standard IPAC education to the context of the COVID-19 pandemic and outbreaks. | *‘The education around the change in direction and general IPAC measures is time consuming.’ ‘The documentation and audits are time consuming.’*  *‘People are not retaining the information or applying it consistently.’ (P1018)*  *‘Lots of educational needs for the staff during the outbreak, they thought that they had a good understanding of what they needed at the time but there were a lot of PPE practices or general knowledge where there was misunderstanding and miseducation about some things from the media.’ (P1008)* |
| Staff did not implement IPAC guidance consistently | Sites experienced inconsistent implementation of IPAC protocols from their staff, such as not always social distancing, not maintaining consistent hand hygiene, and irregularity with how staff wore PPE. | *‘We are struggling with consistently maintaining IPAC precautions like cleaning and distancing. People relax over time and we can’t allow them to relax. It is a struggle to get the point across gently.’ (P1018)*  *‘And then compliance, with making sure they’re wearing their PPE. It’s been a huge factor. They determined we should wear goggles at all times, as well as other IPAC criteria. Buy-in from staff has been difficult. We notice a lot of times on the unit, we notice that goggles are not worn, or worn on the top of their heads.’ (P1022)* |
| Residents with limited capacity were not able to follow IPAC protocols | Staff have experienced challenges with ensuring residents with dementia are following IPAC protocols. There were challenges around keeping PPE on residents with dementia, ensuring that they maintained hygiene (i.e., handwashing), and maintaining isolation of ill residents. | *‘They cannot keep it on. Our residents are more in the advanced stages, you cannot put PPE on them, you cannot confine them to their rooms…So we had to isolate those who didn’t. It was reversed isolation- those that were negative were isolated.’ (P1214)*  *‘In my observation we have residents that are dealing with dementia and agitation, we have one floor where we have residents wandering all the time… Hard to do isolation because residents won’t stay in their rooms… Those are the challenges for me would be managing IPAC while managing those residents safely…For example changing the linens after toileting a resident and making sure they’re washing their hands properly.’ (P1029)* |
| Homes had a hard time keeping up with rapidly evolving protocols and communicating changes to families/caregivers and staff, which led to confusion, fear, anxiety and anger | Directions and protocols were constantly changing, making it difficult for staff to keep track of current protocols and practices. This was especially true for RH who did not always have tailored public health guidance available to them. Many sites had difficulty keeping up with the updates as a result, and experienced challenges with effectively communicating the ongoing changes in IPAC protocol to staff, residents, and caregivers/families. | *‘There has been so much information and so many changes in information, it’s almost information overload, each week something changes. The uptake is not as fast and other things move quickly. Keeping up with changes and how to message to staff here’s what we used to do vs here is what we need to do now.’ (P1016)*  *‘Every time we think we’re – oh okay right now we are doing this but a week later it changes to something else. It is hard to keep up with.’ (P1206)*  *‘The ever-changing directives, conflicting, nuanced directives, and the institutionalization of the directives have had a tremendously negative impact on the Retirement sector. For residents coming in it has resulted in deconditioning, fear, anxiety, mental health and depression and that’s real. Every surge has been different, and so what we learned in April 2020 is no longer the same environment even though it’s still within the pandemic cycle.’ (P1217)* |
| Resources shortages, including PPE and access to COVID-19 rapid tests | Sites experienced shortage or poor access to PPE for staff members, making it difficult to follow IPAC protocols. This was particularly an issue during the first wave, where there were shortages on all PPE (which they were able to resolve over time), but was also seen later in the pandemic during a shortage of N95 masks following guidance changes.  Following recommendations to employ rapid COVID-19 testing during the Omicron wave, sites experienced challenges in acquiring sufficient rapid COVID-19 testing kits in a timely manner, resulting in shortages. | *‘When the pandemic hit, we were quite unprepared, they did not have any pandemic supplies. When we went into their pandemic room they saw that everything expires, we were short of PPE, they had staff shortages. It took us about 6 months to reach equilibrium. This is definitely resolved now, we have sufficient amount of supplies.’ (P1021)*  *‘The team is very good with wearing PPE; we’re struggling right now with getting N95 or KN95, it’s a huge struggle. So, we’re worried about that right now.’ (P1215)*  *‘Access to rapid tests is difficult; supposed to arrive in 5 days but it takes 2 weeks.’ (P1213)* |
| Lack of funding to cover costs related to N95 mask fit testing, staff training for IPAC protocols | While some homes had enough funding or resources in-house to support IPAC (e.g., N95 mask fitting; IPAC protocol training), other homes did not have the necessary resources in house nor sufficient funding to do so. | *‘To bring in the mask testing. We’re behind on that because we hired a lot staff since she was in. when she was here we didn’t have a lot of time but she’s willing to work with each of the staff members so that she can properly show how to wear mask, why they wear a mask, when to change their masks. She’s willing to do all of that with them but she was being paid by the hour I only had so much money I could put towards that.’ (P1212)*  *‘For a home of my size, to get someone to come in is a couple thousand dollars. So it is privately paid and the other thing I do is I take on people that can’t even afford. So affordability for those extra things is a challenge.’ (P1214)* |
| Physical environment of homes not conducive to IPAC implementation | Homes experienced challenges around finding physical space for IPAC supplies, disinfecting large areas, and ensuring physical distancing in an environment that was not built to handle such expectations (e.g., lack of space, small physical space). | *‘The physical layout of the building means it’s not always easy for people to engage in social distance when having meals and such for break time for staff.’ (P1024)*  *‘One of the difficulties during outbreak is space. We are an older building where they were never meant to have a pandemic in. We lack division of units with fire doors. We had to change their sections to be able to make for a safer IPAC noted sections. We had to get creative with different spaces to store PPE.’ (P1008)* |
| PPE fatigue | Staff members and caregivers became tired of wearing PPE, due to discomfort that comes from wearing many layers of PPE consistently throughout their shift; this worsened in warmer weather. This led to PPE burnout and decreased compliance to PPE in many instances. | *‘If a unit goes on suspect outbreak it is difficult for staff – they need to wear PPE and change every time they go into a different room… but when they do audits they see that not all of them are complying – they will hear that they are tired with unit on suspect outbreak, they are tired – there is lots of PPE fatigue… Still not 100% compliant due to fatigue.’ (P1035)*  *‘Staff complained wearing mask and face shield, they do complain… You really cannot blame--it’s been so long wearing it, the new rules came out that our screeners have to wear N95, full PPE and face shield… N95 there is no way you can make comfortable because it has to fit very tightly so nothing goes in… Staff are adhering to it but no one likes it.’ (P1031)*  *‘In the summertime – PPE fatigue is dependent on the weather. In the summertime, staff were really struggling. It was really hard for them to wear the full face shield and provide care to residents – you are really sweating under there. Especially in the shower. Now that is not a problem – if you are vaccinated you now do not need to wear the face shield.’ (P1033)* |
| Family pushback on IPAC protocols | The pandemic led to new levels of distrust and pushback from caregivers/family members to loosen restrictions. Family members called to complain about not being allowed to visit. Some family members requested residents be cared for by vaccinated staff members, creating tensions between staff. |  |
| Fears of returning to normal and loosening IPAC restrictions | Some found adapting to the earlier stages of the pandemic easier because the focus was on constant change. Now some are finding it difficult to loosen IPAC restrictions, for fear of another outbreak. Some expressed a fear of “returning to normal”. | *‘As this has worn off the other needs are ramping back up.’*  *‘You have demands for other things. I just feel like we have been hanging on for dear life for a year and a bit and now you’re telling me to let go? It’s hard to let so many people come in after we have been so cautious.’*  *‘There has been a disconnect to how tightly we have been holding on to things and now letting go.’ (P1025)*  *‘We have a fear of going back to normal; we have been very very careful and I think that’s why we haven’t had a second outbreak or have things like that because we’re watching all the breaches that we can’t control happening. We’re at least being very safe in here.’ (P1206)* |
| **Supports and Strategies Implemented by LTCH and RH to address IPAC challenges** | | |
| **Theme** | **Description of Theme** | **Quote** |
| Having a dedicated IPAC manager/nurse/champion | IPAC representatives helped provide advice, answer questions, and supported each site and their staff to ensure IPAC protocols were properly followed. | *‘IPAC team – biggest support during the past year – with [hospital]… they provided very good concrete advice – giving other pointers about this is what they do, this is what the suggestion is, acknowledging that people have lives outside of the RH and acknowledging this and the nervousness of how do you go home and protect your family.’ (P1207)*  *‘There has been some non-compliance with staff and visitors but we have recently implemented IPAC Staff Champions, so they give feedback to us as to who they identified as not following guidelines. So that is helpful. We try to educate the staff when we have our weekly meetings so we have someone show us how they would do their PPE, like donning and doffing, and educate them on the role of the IPAC Champion. So because they are really educating each other rather than us educating them it is like we have a team working alongside the management to ensure IPAC precautions are upheld.’ (P1217)* |
| Consistent communication with public health units | Some sites had regular communication with their local public health units that provided them with support, education, and information. | *‘They have regular calls (biweekly) with their local public health [unit] – has been a great forum to share and exchange ideas with other homes, ask questions when they are unclear, etc. – there has been so many changes almost daily – that platform has been really great.’ (P1211)*  *‘When there are cases related to IPAC (i.e., COVID cases) this is the only time they get in touch with [hospital] and [public health units]… they keep in consistent communication about any potential IPAC issues.’ (P1035)* |
| External supports from hospitals or PHU (IPAC guidance, physical or financial resources) | Sites received a range of support/ resources from local public health agencies and external organizations/ hospitals. There was also financial support (e.g., from provincial government) to RH to purchase PPE and cleaning supplies, and hire emergency services to provide onsite support. | *‘The support from the province, I think it comes from the top, we were given funding to acquire PPE and the cleaning stuff so that helped the staff to be comfortable and also the residents.’ (P1214)*  *‘During first outbreak they got support from [local health integrated network] LHIN – got redeployed nurses to support them, they also got deployed some staff from [hospital].’ (P1012)* |
| Use of multi-pronged strategies to disseminate IPAC updates to staff | Sites utilized a variety of strategies to disseminate IPAC information and updates to their staff, tailoring formats or leveraging communication strategies they already had in place.  Strategies included huddles, town halls, emails, calls, handouts, bulletins, in combination with innovative, interactive, and informal approaches to sharing information. | *‘We send weekly reminders to staff, through school messengers and emails. We talk about PPE and things to stay on top of. A lot of signage in the building too. And in the outbreak we were doing almost daily announcements. Daily through the phone system, through the email system. And daily things like reminders and tips to do…we also did IPAC huddles, so safety concerns, at the beginning of the shift, what are the challenges, what’s not working? Ok let’s go fix that for you.’ (P1020)*  *‘Small group in service or team huddles where there’s a lot of interaction… I try to run my programs where people can contribute their experiences and its very informal… I think the staff respond well to the interactive aspect.’ (P1029)* |
| Monitoring and audits on IPAC compliance | Sites monitored and conducted audits to ensure staff, caregivers, and visitors comply with IPAC protocols. | *‘We’ve done a lot of audits, we have external parties coming in for assessment, we follow the IPAC checklist.’ (P1023)*  *‘What they’ve been doing on PPE is daily manager walkabouts – daily the managers would go around witnessing staff doing IPAC protocols and would correct them on the spot.’ (P1013)* |
| Having leaders who are committed to transparency with staff, families/caregivers | Commitment to communication from leadership, and transparent, ongoing communication among staff, and between staff and caregivers/ families were effective; included open dialogue on challenges around adherence to IPAC guidelines.  This fostered trust. | *‘During height of COVID, this was [on a] daily basis… I go everyday on each floor. throughout the shifts, the staff are always calling me asking me questions, the communication channel is always open to anyone we are already available 24/7 we are always available to our staff. For family members I will email them or phone them to inform them, what’s new what changes are coming.’ (P1031)* |
| Homes with physical space conducive to IPAC measure implementation | Having a larger sized home facilitated following IPAC protocols on physical distancing and social isolation. Sites also leveraged other techniques, including restricting movement through cohorting. | *‘This is due to the mere size of the home and the number of people coming in. Being such a large home allowed them to physically distance in a way that is able to help them with their IPAC practices.’ (P1033)*  *‘When we were on outbreak each floor had a designated break room so we were cohorting staff to the floors.’ (P1009)*  *‘Our homes were certainly one of the lucky ones where we had space so we could change one of the rooms for visiting, other homes are right in the middle of the city that have no real estate so our community has been wonderful we’ve been grateful for that.’ (P1206)* |
| Having leaders with experience navigating public health emergencies | Sites with staff who either worked through previous pandemics or earlier COVID-19 waves, were able to help sites navigate the pandemic and associated outbreaks. | *‘One benefit was our [director of care] DOC...before. She had experienced SARS so she implemented a lot of things she had experienced from SARS at the home. Having that veteran, who know what to implement quickly, helped us feel safe and more prepared in the middle of a lot of people around us not knowing what to do. That was a big factor in our success.’ (P1020)*  *‘Because of their experiences then during the first wave, they were able to build the IPAC program to how it is now. That’s how they came to creating the IPAC role, they realized they need one person to manage this. They are still in and out of outbreak but with this role in place (IPAC person) they are able to manage it better in the third wave now compared to before.’ (P1012)* |

Table 2: IPAC Challenges and Strategies Implemented
