## Supplemental Table 3 for "Challenges facing Canadian Long-Term Care Homes and Retirement Homes during the COVID-19 pandemic"

| **COVID-19 Vaccine Challenges Experienced by LTCH/RH During the COVID-19 Pandemic** | | |
| --- | --- | --- |
| **Theme** | **Description of Theme** | **Quote** |
| Lack of vaccine availability | Homes experienced challenges with COVID-19 vaccine availability due to interim supply chain issues (i.e., Canada-wide 2^nd^ dose delays) – this delayed vaccination of their staff. | *‘All set to roll something out in February, then the vaccine supply dried up; Biggest challenge right now is not being able to run a few vaccine clinics maybe a week apart (from each other), potentially get our vaccination rate up for first dose for staff; It’s hard to promotion and whip up enthusiasm and then not being able to produce the vaccine.’ (P1013)*  *‘There is a back order on vaccine, I think it’s everywhere right now. The day before yesterday I received the vaccine order that I ordered two weeks ago. If I had more vials right now I would give more because lots of staff are writing up, giving their name everyday asking for it.’ (P1031)* |
| Mistrust around vaccine safety | There was mistrust from staff, caregivers, and residents around vaccine safety due to the following reasons:   - Rapid vaccine approval timelines - Changes in vaccine schedule - Fears about long-term effects and vaccine safety and particularly around fertility, breastfeeding, and pregnancy   Members of marginalized communities also experienced mistrust rooted in systemic and historical mistreatment by government and health institutions. | *‘… their biggest complaint is, [the studies on vaccines] It’s on a piece of paper. It looks like someone typed it on word, so how do they know the logistics? Case studies? They want more credible info on these vaccines. I myself wanted that as well. So those kinds of resources, with the backing of how the study was done, how vaccine was created, would definitely help percentages go up.’ (P1022)*  *‘Some don’t know the long term effects of it and that’s the problem. It’s just the unknown… And over the internet they read certain things like a year or in two years we’re all going to die from it. And I think they’re talking about the booster, and the issues with AstraZeneca and before you couldn’t mix then and now you can. And if you took the AstraZeneca you can’t travel. And the Johnson and Johnson that was bad thing too. Adding to the unsureness.’ (P1027)*  *‘Some females think it might affect their reproductive system and they don’t want to get vaccinated… some young women are thinking of getting pregnant in the future and it’s a lot of the unknown.’ (P1027)*  *‘Staff population is largely Black community, they have a lot hesitancy around previous vaccinations, Black people have been used as guinea pigs in the past for vaccines; there was a cultural vaccine hesitancy, they have a very diverse staff so certain cultural groups were very hesitant.’ (P1013)* |
| Belief that the vaccine, particularly boosters, will not improve or impact health outcomes | Some staff did not believe the impact would improve health outcomes; this sentiment was further driven by positive COVID-19 cases and continued COVID-19 restrictions after vaccination. | *‘Some think they got COVID already so they are already immune and don’t need vaccine.’ (P1012)*  *‘They just started doing 3 doses for residents. This has surprisingly been a challenge – people got 2 but they feel like why should I be getting another 1. Shouldn’t I have antibodies?’ (P1033)* |
| Lack of knowledge, access or ability to get to a vaccine clinic | Staff at sites experienced challenges with accessing vaccine clinics, some of which were due to the following:   - Lack of transportation to the vaccine clinic - Inability to find local vaccine clinics - No protected time to travel to a vaccine clinic (staff would rather wait till the vaccines are available at the site) - Inability to book an appointment following the new online vaccine clinic booking system; compounded with long waitlist times | *‘It is so difficult to find a place to get vaccinations though…I was saying how I am encouraging my staff to get vaccinated but am having trouble finding a place.’ (P1214)*  *‘… I got an email they’re opening a clinic at 3 locations…But most of my team are like we can’t even get through. And the appointments they did have booked, some of them had appointments booked in March, so in order to try to book an appointment through this portal, then they had to cancel their other appointment so now they lost their other appointment. So it’s not set up really well… We’ve been very fortunate here because they’re [staff] like “yeah I want to get it”. It’s just a matter of being able to get it.’ (P1215)*  *‘Some of them [staff], I can’t say how many, but definitely transportation is a barrier for staff. Another barrier is time, people wanting to wait until it’s available in the home but it’s not even on the horizon yet.’ (P1015)* |
| Families’ concerns about vaccine safety impact vaccine uptake among residents who do not have capacity to provide consent | Families who did not want to get vaccinated prevented their loved ones from receiving the vaccines. | *‘Some family that chose not to get the vaccine won’t let their loved ones in residence get it either; the ones that have chosen not to get it they are pretty adamant on their decision and not letting their loved ones get it.’ (P1012)* |
| Beliefs that vaccine mandates are an infringement on labour laws, personal liberties | Sites experienced staff pushing back against vaccine mandates, voicing their concerns that it was against their rights and that they were being forced by their organization to get the vaccine. | *‘Now province is putting in mandate that by Sept 30 everyone needs to have first dose – getting feedback that it is against their rights, they need exception for X reason (e.g., religious beliefs) – this is the biggest challenge.’ (P1033)*  *‘A big change is, now they say, if you want to travel, vacation, you have to get it. It makes people unfortunately feel that they have no choice.’ (P1020)* |
| **Supports and Strategies Implemented by LTCH and RH to address COVID-19 Vaccine Challenges** | | |
| **Theme** | **Description of Theme** | **Quote** |
| Individual-level strategies to promote vaccine confidence | Sites had success with targeting each staff member at an individual-level and engaging leadership to educate staff on the vaccine, promote uptake, and provide them with access to evidence-based information. | *‘Our medical director is always available if any staff have any questions about the vaccines, so the staff can set up an appointment to meet with them any time.’ (P1031)*  *‘And if there was someone who was hesitant, we would address it 1:1, and if it escalated then the manager would talk with the individual saying, “it is all good, here’s all the research on it”, and if it didn’t work then the owner/operator would step in and by then almost all the staff got it. We had a few that lingered because they wanted to talk with their doctors, but we were able to debunk myths and I think taking that management strategy had a big impact.’ (P1217)*  *‘They finally have a vaccine clinic on site for residents; Once they had a clinic on site with people they knew (e.g. medical director to speak with staff), that helped with vaccine hesitancy because people they are familiar with.’ (P1013)* |
| Organizational level strategies to promote vaccine confidence | Sites also ensured that staff were provided with evidence-based information and education about the vaccine to combat misinformation, including information to explain the rapid approval process. Sites found that informational town halls were an effective strategy for increasing vaccine uptake.  Sites utilized opinion leaders and vaccine champions to promote vaccine uptake. Strategies ranged from bringing in staff to speak to historically marginalized populations, having management being the first to get vaccinated, holding vaccine campaigns with champions, and bringing in team members who were negatively impacted by COVID-19 to speak about their experiences. | *‘During the early stage of vaccination, staff were having concerns about side effects – they were able to provide education about the vaccines, were able to create a pamphlet with FAQs, ingredients, safety for pregnancy and age.’ (P1035)*  *‘… we had management rolling up their sleeves and saying, “I am doing it, I rolled up my sleeves”. We had our DOC go first, the hospital affiliated with us at the time sent busses, and with each bus 1 manager went and got the vaccine.’ (P1217)*  *‘IPAC lead, from Jamaica from Humber River – she had gone around to various cultural groups and speak to them which has helped for vaccine hesitancy.’ (P1013)* |
| Incentives to support vaccine uptake | Some sites provided resources to staff to be vaccinated. This included paying transportation and parking fees for staff as an incentive. | *‘Some of them, I can’t say how many, but definitely transportation is a barrier for staff. So we’ve reached out to some and say we are willing to pay taxi and uber, and we have one more staff sign up because of that.’ (P1015)*  *‘We paid staff to go, even for transportation and parking, if they went to get vaccinated at the hospital site. That was encouragement for them to go, doing it outside of working hours, for them to go.’ (P1023)*  *‘And we also would pay our staff for 3 hours of work if they chose to get vaccinated on their own time, so that was an incentive.’ (P1217)* |
| Improving ease of access to vaccinations for LTCH/RH populations | Sites’ connection to IPAC supports including IPAC hubs facilitated access to vaccine supply and appointments. Sites were also able to overcome logistical barriers to vaccine access by supporting staff with appointment booking or by leveraging mobile clinics/on-site clinics; this likewise included offering vaccines to caregivers while they were onsite visiting residents to facilitate/ encourage uptake. | *‘There are some people she was able to assist with making the appointment for them on her computer because don’t know if technology is a barrier. Got about 4/5 people to get the vaccine from doing that.’ (P1008)*  *‘They finally have a vaccine clinic on site for residents; Once they had a clinic on site with people they knew (e.g. medical director to speak with staff), that helped with vaccine hesitancy because people they are familiar with -- This Friday through [Public Health Unit], we’ve been given the opportunity to vaccinate with Pfizer on-site; A few staff members put together this vaccine clinic; Now there is a growing list of people waiting to get vaccinated.’ (P1013)*  *‘The staff give priority to the essential care givers for vaccine – since most of the residents are in the 90s and their children are in the 70s, they are at higher risk; they had a mobile clinic come to the home – 100% residents are vaccinated.’ (P1201)* |

Table 3: COVID-19 Vaccine Challenges
