## Supplemental Table 4 for "Challenges facing Canadian Long-Term Care Homes and Retirement Homes during the COVID-19 pandemic"

| **Well-being Challenges Experienced by LTCH/RH During the COVID-19 Pandemic** | | |
| --- | --- | --- |
| **Theme** | **Description of Theme** | **Quote** |
| LTCH staff experienced burnout, moral injury, PTSD, stress,  health challenges and general lack of well-being and morale during the pandemic | The pandemic negatively impacted the mental health of staff. Challenges to well-being included working long hours (consistent overtime, no vacation), inability to care for residents in the way they wanted to due to IPAC requirements and staff shortages, working alongside agency staff who were unknown to home staff [and unfamiliar with the home/residents], high levels of stress and always being on alert, fear of bringing COVID-19 to the home, and witnessing the deaths of residents. | *‘Our staff our tired, burnt out, it’s just wellness and supporting them, they’ve been through a lot, as I said with our clientele, a lot of people don’t have families so we become their family. I think staff might be burnt out.’ (P1009)*  *‘Staff are concerned that they are continuing to have outbreaks; concerned about themselves and their families; One of the big things is mental health, a lot of staff are extremely stressed and burned out; A lot of them were here from the beginning and watching the residents die and coworkers get sick; Even though it’s better than last year it’s still hard.’ (P1012)* |
| Staff generally reported a lack of access to appropriate mental health and well-being supports in homes | Many homes did not have appropriate mental health supports available. As the pandemic progressed, some supports that were initially put in place were discontinued due to limited resources. | *‘Supporting staff wellness-- there’s not a lot that we do right now, we have paid sick days for staff , but in terms of more proactive things that are tools for personal mental health and wellness we don’t do a lot.’ (P1029)*  *‘Morale is low with the staff – before (early 2021) management was providing free meals, snacks for the staff during COVID – it was taken away during the 2nd wave – staff are not really getting any incentives. No EAP [employee assistance program] for staff as well.’ (P1035)*  *‘Right now all the programs are limited, how do we make sure that people still have their social needs met when its limited- they don’t have the human resources to support every resident and staff with their needs.’ (P1201)* |
| When available, staff did not access EAP and other well-being resources; stigma was perceived as a factor. | In situations where well-being resources and supports existed, there remained a lack of uptake. Staff did not have capacity to engage with them, and/or had concerns about stigma and privacy. | *‘There is still stigma around mental health and don’t want people to think they are crazy if they reach out to help; They are reluctant because they feel like there is a judging factor; They might not want to share which RH they came from but it’s just about knowing that there’s other people that went through the same circumstances.’ (P1209)* |
| **Supports and Strategies to address Wellness Challenges** | | |
| **Theme** | **Description of Theme** | **Quote** |
| Some home leaders implemented a variety of strategies to address well-being | To support staff and resident wellness, sites implemented diverse strategies, which included:   - Providing behavioural supports for residents through Behavioural Supports Ontario (BSO) - Limiting number of staff shifts and encouraging use of vacation time - Leveraging support from external organizations to promote staff wellness (e.g., wellness resources from Local Health Integration Networks (LHIN) and regional hospital networks; drop in counselors or psychogeriatric nurses) - Having clinical staff on site with expertise in promoting wellness (e.g., social worker, occupational health) - Offering Employee Assistance Programs - Hosting social activities and staff appreciation events, providing gift certificates - Distributing well-being resources (e.g., bulletins, self-serve resource table) | *‘Yes, got a BSO social worker to stay in home. Social worker. Goes above and beyond and is really their go to. Helps with rec – fun activities and ideas. She helps with everything.’ (P1006)*  *‘They are so dedicated to make sure they are taking care of their residents and staff, this year we have started the vacation planning early in the year and made it clear to them to do whatever they can to make sure they have their time off; This has been helpful in making sure they don’t have burnout and ensuring time off.’ (P1021)*  *‘We have a team that they can call anytime, Homewood health, and there are counsellors that can help them through.’ (P1209)*  *‘One of our managers is implementing a wellness program in our home that’s just focused on wellness among staff. Currently only have a “wellness committee” being created, to encourage other staff from other departments to participate in it. That’s also a work in progress; it’s not just nursing staff, they want PSWs, dietary aids, physio aides, etc. They’re creating posters, etc. to get them to join in.’ (P1009)* |

Table 4: Well-being Challenges and Implemented Strategies
