## Appendix B for "Challenges facing Canadian Long-Term Care Homes and Retirement Homes during the COVID-19 pandemic"

### **Wellness Hub Needs Assessment**

### Please note that the interview guide is semi-structured and may be tailored slightly to probe for specific challenges experienced by the LTCH/RH and/or to react to specific COVID-19 circumstances (e.g., third wave).

| **Question** | **Notes** |
| --- | --- |
| **Section I: Background information**  **Time: 5 minutes** | |
| 1. Please describe your professional role(s) and responsibilities in your LTCH/RH setting(s). |  |
| 2. What has your personal experience, as well as the experience of you LTCH/RH setting(s), been through the COVID-19 pandemic to date?    a. *(For site-level interviews)* Is/are your setting(s) experiencing or has/have your setting(s) experienced a COVID-19 outbreak?   *[If yes]*   i. How many outbreaks have your setting(s) experienced? ii. What is/was the magnitude of these outbreaks (i.e., small, medium, large – no need to specify numbers)? iii. Is/was the outbreak focused in residents or staff or both? |  |
| **Section II: Challenges and opportunities during COVID-19**  **Time: 10 minutes** | |
| 3. What are some of the main challenges that you/your co-workers/your LTCH/RH setting(s) have experienced throughout the COVID-19 pandemic to date?    a. What, if any, are some challenges that you experienced relating to:       i. Infection prevention and control       ii. Resident programs       iii. Staff wellness       iv. Vaccine confidence       v. Staff supports (e.g., wraparound care such as supports to effectively quarantine)       vi. Staff decision making       vii. Other   *Probes for each challenge shared:*   b. What factors, if any, do you think contributed to this challenge (e.g., supply and personnel needs, government support, etc.)? c. At what stage(s) of your COVID response did you find this challenge (i.e., prevention, outbreak management, or recovery post-outbreak)? d. Did your LTCH/RH setting(s) implement any changes to practices and policies to address these challenges?    i. Could you expand on what made these changes particularly helpful for you/in your setting(s)?    ii. What changes did you feel were missing or lacking, but would have been helpful? e. What additional changes, if any, at the individual, organizational, and policy level do you think could help mitigate these challenges? f. Are there any organizations that you trust and either work/have worked with or accessed resources and other supports from? |  |
| 4. *(For site-level interviews)* What supports at the individual, organizational, and/or policy level have you/your co-workers/your LTCH/RH setting(s) found helpful during the COVID-19 pandemic, if any?    a. Could you expand on what made these factors particularly helpful for you/in your setting(s)?    b. At what stage of your COVID response did you find this factor helpful (i.e., prevention, outbreak management, or recovery post-outbreak)?    c. What additional factors, if any, at the individual, organizational, and policy level do you think would have been helpful? |  |
| **Section III: Previous, current, and anticipated support needs during COVID-19**  **Time: 10 minutes** | |
| 5. What, if any, are **(1)** your current support needs on an individual-level, **(2)** in your perspective, the current support needs of your co-workers, and/or **(3)** your LTCH/RH setting(s)’s current support needs?    a. What specific resources (i.e., personnel, funds, information, tools, etc.) do you think may help address these support needs?    b. *(For site-level interviews)* How do you/does your setting prefer to receive support for your needs (e.g., through an online platform, through an in-person coach and support staff, through a webinar, etc.)?    c. Are there any other factors that would be important to consider when providing supports to you/your co-workers/your LTCH/RH setting(s)? |  |
| ***Only ask the following two questions if time permits and they have not previously been addressed through question 5:*** | |
| 6. What, if any, supports do you anticipate that you/your co-workers/your setting may need in the future?    a. Why do you think that there may be this future need?    b. What, if anything, could help you prepare for this potential future need? |  |
| 7. Could you please describe the break room set-up in your LTCH/RH?    a. Is there enough room for staff to physically distance during their breaks?    b. Are IPAC protocols (e.g., wearing masks except when eating, physical distance, hand washing) being followed in the break rooms?       i. What do you think are some challenges to having IPAC protocols followed in break rooms?       ii. What are some facilitators to having IPAC protocols followed in break rooms? |  |
| 8. What, if any, supports did you/your co-workers/your setting(s) previously need but are no longer in need of?    a. Why have your needs changed (i.e., did you access these supports or have your needs changed, or both)?    b. *[If they accessed supports]* What supports did you find particularly helpful?       i. What, if anything, did you like and/or dislike about how you received these supports (i.e., the modality, timelines, etc.)? |  |
| **Section IV: Wrap-up**  **Time: 2 minutes** | |
| 9. Are there any other challenges, opportunities, or support needs that you/your co-workers/your LTCH/RH setting(s) have experienced or are currently experiencing during the COVID-19 pandemic that you would like to share? |  |
| 10. *(For site-level interviews)* As part of your participation in the IPAC+ study, you may be asked to temporarily store data (e.g., dried blood spot samples, demographic questionnaires) in a secure space in the LTCH/RH.     a. Do you have a space available that could be used for this?     b. Is there anything that we can do to support with creating a secure space designated for this? 11. *(For site-level interviews)* How would the saliva testing protocol work best with your setting? Do you have a place for on-site testing of should these samples be done at home for symptomatic individuals or those with high-risk contact? |  |
